# Decreased Awareness of Cognitive Decline is Associated with Multimodal Alzheimer’s Disease Biomarkers in Cognitively Unimpaired Individuals

**DOI:** 10.64898/2026.03.03.26347515

**Authors:** David López-Martos, Marc Suárez-Calvet, Gemma Salvadó, Raffaele Cacciaglia, Mahnaz Shekari, Armand González-Escalante, Andrea Horta-Barba, Helena Palma-Gudiel, Marta Milà-Alomà, Anna Brugulat-Serrat, Carolina Minguillon, Matteo Tonietto, Edilio Borroni, Gregory Klein, Clara Quijano-Rubio, Gwendlyn Kollmorgen, Henrik Zetterberg, Kaj Blennow, Juan Domingo Gispert, Gonzalo Sánchez-Benavides, Oriol Grau-Rivera, the ALFA study

## Abstract

**INTRODUCTION:** Alzheimer’s disease (AD) diagnostic guidelines emphasize subjective cognitive decline (SCD) preceding mild cognitive impairment (MCI), implicitly assuming awareness of cognitive decline (ACD) is preserved in preclinical AD. This study aimed to evaluate associations of decreased ACD with multimodal core AD biomarkers in cognitively unimpaired (CU) individuals.

**METHODS:** We analyzed data from CU individuals with baseline CSF biomarkers and 3-year longitudinal neuropsychological assessment (ALFA+ cohort). Decreased ACD was defined by concurrent decline in episodic memory and awareness using robust longitudinal references (Free and Cued Selective Reminding Test, Memory Binding Test, Wechsler Memory Scale, and Subjective Cognitive Decline Questionnaire). Biomarker outcomes included plasma and CSF p-tau181, p-tau181/Aβ42, p-tau217; Aβ ([¹⁸F]flutemetamol) and tau PET ([¹⁸F]RO948). Associations of ACD with AD biomarkers were evaluated using linear regression models. Sensitivity analyses were restricted to individuals with memory decline.

**RESULTS:** 350 CU individuals were included (mean age 61 years; 60% female; mean education 14 years; 35% CSF Aβ-positive). Episodic memory decline was identified in 61 (17%) individuals, of whom 25 (41%) also exhibited awareness decline; meeting criteria for decreased ACD. This group demonstrated greater levels of AD pathology compared to the remaining sample. Among fluid biomarkers, CSF p-tau217 showed the strongest association. Neuroimaging revealed elevated frontoparietal Aβ PET, alongside temporal, insular, and frontal tau PET deposition. Sensitivity analyses showed that, at the same threshold of memory decline, decreased ACD reflects greater AD pathology.

**DISCUSSION:** Standardized assessment of cognitive awareness, integrating objective neuropsychological performance with subjective reports, may provide a crucial extension of current clinical frameworks.

## Introduction

A substantial body of research has focused on subjective cognitive decline (SCD) in the preclinical stage of Alzheimer’s disease (AD), defined as self-perceived worsening of cognitive function in the absence of objective cognitive impairment.^1–3^ SCD has been incorporated into diagnostic frameworks, ^4,5^ and it is widely considered an early stage preceding mild cognitive impairment (MCI). However, cognitive complaints are common in aging, reported by 50–80% of adults over 75 years, are influenced by mood, personality, and psychosocial factors, and may reflect diverse medical conditions.^1,2^ Consequently, SCD constitutes a non-specific predictor of clinical progression in AD,^6,7^ although its prognostic value is substantially enhanced when it is considered in conjunction with biomarker evidence of AD pathology.^8^

In this context, the distinction between self-reported and informant-reported SCD has received little systematic attention, despite these measures carrying fundamentally separate prognostic implications; with informant-reported SCD being a more reliable predictor of future cognitive decline and dementia risk.^7,9,10^ Reports from study partners or informants have provided a crucial complementary source of information, offering an external perspective on cognitive and functional changes that may otherwise go unnoticed by the individual or patient themselves, as progression of AD is associated with decreased insight into cognitive performance.^10–13^

Most research has placed substantial weight on SCD without explicitly recognizing this limitation, implicitly assuming that cognitive awareness is preserved across the early AD *continuum*.^8^ Emerging evidence challenges this assumption, showing that cognitive awareness may decline earlier than previously recognized, progressing with underlying AD pathology across the preclinical stage.^14^ This raises a critical question: if self-awareness changes in the preclinical stage of AD, how can self-reported SCD alone reliably identify individuals at risk of progression?^1,13^

Awareness of cognitive decline (ACD), a construct integrating both objective and subjective metrics, provides a critical extension of current AD clinical frameworks.^8^ ACD can be determined by the discrepancy between objective neuropsychological performance and subjective reports of cognitive decline. However, systematic evaluation of ACD in relation to AD biomarkers remains limited in CU populations, as most research has been conducted in MCI and dementia stages, in the context of overt clinical symtpoms.^15^ This gap in the literature is particularly important given the need to characterize the longitudinal trajectory of ACD in the preclinical stage of AD.^8^

Studies on amnestic MCI patients with anosognosia, defined as impaired cognitive awareness, have provided key insights into how high-order cognitive processes, such as self-awareness and meta-cognition, are affected due to AD pathological processes, reflecting a key discrepancy between episodic memory impairment and the subjective perception of actual performance.^16,17^ Crucially, this pattern may already be detectable in subclinical forms during the preclinical stage of AD, where ACD begins to change alongside subtle cognitive decline, and this may encompass both increased and decreased ACD within the cognitively unimpaired (CU) range. ^18,19^

Among these subtle changes, decreased ACD is of particular concern, as it may represent a subclinical manifestation of anosognosia, preceding formal diagnostic thresholds.^20,21^ The prevalence estimates of anosognosia ranging from 20% to 80% across MCI and AD dementia stages,^22,23^ which vary by diagnostic criteria and increase with disease progression, suggest that subclinical progression of decreased ACD occurs longitudinally before overt anosognosia emerges in symptomatic stages. This highlights why exclusive reliance on self-reported SCD may overlook CU individuals with reduced insight into their cognitive decline.^24–27^ Such individuals may fail to report cognitive complaints or seek medical help despite accumulating AD pathology, a phenomenon well-established in later disease stages but potentially present much earlier.^28–31^

However, this is not explicitly acknowledged in current diagnostic and staging frameworks, and this lack of recognition represents a significant vulnerability.^4,5^ Specifically, liberal use of SCD criteria may fail to capture clinically meaningful heterogeneity associated with decreased ACD in preclinical AD, which is analogous to systematic diagnostic errors previously documented in MCI.^32–34^ Altogether, these limitations have important implications for early detection and risk stratification in AD.

To address this gap, the present study determined ACD using a robust longitudinal normative neuropsychological framework, aiming to evaluate its association with multimodal core AD biomarkers, directly examining the clinical and biological relevance of decreased ACD in CU individuals at increased risk of AD dementia.

## Methods

### Study Participants

This study was conducted within the Alzheimer’s and Families+ (ALFA+) cohort, a longitudinal extension of the ALFA cohort at the Barcelonaβeta Brain Research Center, Barcelona, Spain.^35^ The ALFA cohort recruited CU individuals aged 45–74 years, free from major medical, psychiatric, or neurological conditions. The ALFA+ cohort represents a selected subset of ALFA enriched for AD risk factors, including parental history of AD and carriage of the apolipoprotein E (*APOE*) ε4 allele. Eligibility criteria for ALFA+ included prior participation in ALFA and willingness to undergo clinical and neuropsychological assessment, blood and Cerebrospinal Fluid (CSF) collection, as well as Magnetic Resonance Imaging (MRI). In addition, a subset was also characterized by positron emission tomography (PET). Participants with cognitive impairment, systemic illnesses, or monogenic AD mutation were excluded. Full details are provided in the Supplementary Material.

In this study, we included CU participants from ALFA+ with available baseline CSF biomarkers data and longitudinal neuropsychological assessments (two visits over a 3-year follow-up). Individuals with normal CSF biomarker profiles or classified within the AD *continuum* were retained (n= 350), whereas those with suspected non-AD pathology were excluded (n= 13), according to the AT(N) classification system (see Multimodal Biomarker Panel and Classification System). Selection criteria were consistent with previous work in the ALFA+ cohort, as previously described. ^36^

The ALFA+ study (ALFA-FPM-0311) was approved by the Independent Ethics Committee of Parc de Salut Mar, Barcelona, and is registered on ClinicalTrials.gov (NCT02485730). All participants provided written informed consent, as approved by the Ethics Committee. The study was performed in accordance with the principles of the Declaration of Helsinki.

### Biomarker Measurements

#### Plasma and Cerebrospinal Fluid (CSF)

Participants underwent fasting blood and CSF collection at baseline.^37,38^ Plasma biomarkers included p-tau181, p-tau181/Aβ42, and p-tau217, while CSF biomarkers included Aβ42/40, p-tau181, t-tau, p-tau181/Aβ42, and p-tau217. All biomarkers except plasma and CSF p-tau217 were analyzed at the Clinical Neurochemistry Laboratory, University of Gothenburg, Sweden. Plasma and CSF p-tau217 were measured by Eli Lilly using an in-house Meso Scale Discovery (MSD) assay. CSF Aβ42/40 was quantified with the NeuroToolKit, a panel of prototype electrochemiluminescence Elecsys® immunoassays on fully automated Cobas® e 601 module (Roche Diagnostics International Ltd, Rotkreuz, Switzerland). CSF p-tau181, t-tau, and CSF p-tau181/Aβ42 were measured with electrochemiluminescence immunoassays on Cobas e 601 module. Plasma Aβ42 and p-tau181 were measured with the plasma NeuroToolKit (Roche Diagnostics International Ltd, Rotkreuz, Switzerland) on a Cobas e 601.

#### Positron Emission Tomography (PET)

Participants underwent longitudinal Aβ PET, with a subset of participants also completing tau PET acquisitions at follow-up. Scans were acquired on a Siemens Biograph mCT scanner with cranial CT for attenuation correction. Aβ PET (∼185 MBq [¹⁸F]flutemetamol) was acquired 90–110 min post-injection and tau PET (368.75 MBq ± 13.02 [¹⁸F]RO948) 70–90 min post-injection (20 min, 4 × 5 min frames for both Aβ and tau PET). Images were reconstructed using ordered subset expectation maximization (OSEM) with time-of-flight and point spread function modeling (Aβ: 8 iterations, 21 subsets, 3 mm; tau: 4 iterations, 21 subsets, 4 mm), co-registered to T1-weighted MRI, and spatially normalized to MNI space via DARTEL. Normalized voxel intensities were computed as Standardized Uptake Value Ratios (SUVRs), using the whole cerebellum as the reference for Aβ PET and inferior cerebellum for tau PET. Baseline Aβ PET Centiloid units were quantified using a validated in-house pipeline.^39^ For consistency, Aβ and tau PET images used for voxel-wise analyses were acquired at the same follow-up visit.

#### Multimodal Biomarker Panel and Classification System

According to previous studies in the ALFA+ cohort, the AT(N) classification system was used to define biomarker profiles in accordance with the 2018 NIA-AA research framework.^4^ CSF biomarker classification was determined using previously validated cut-offs within the ALFA+ cohort: Aβ-positivity (A+) was defined by Aβ42/40 ratio < 0.071, phosphorylated tau-positivity (T_1_+) by p-tau181 > 24 pg/mL, and neurodegeneration-positivity ([N]+) by t-tau > 300 pg/mL.^37^ In line with the 2024 Alzheimer’s Association revised criteria, this study focused on core AD biomarkers.^5^

### Clinical Measurements

#### Tests, Questionnaires, and Measurements

##### Free and Cued Selective Reminding Test (FCSRT)

Episodic memory was assessed using the Spanish version of the FCSRT, which employs a controlled learning procedure.^40^ Participants were asked to learn 16 words presented with semantic cues and completed three recall trials, each preceded by a 20-second subtraction task. Trials included free recall followed by cued recall for unrecalled items, with selective reminding applied only in the first two trials. Delayed free and cued recall was assessed after 25–35 minutes. Variables of interest included total free immediate recall (0–48), total immediate recall (0–48), total free delayed recall (0–16), and total delayed recall (0–16).

##### Memory Binding Test (MBT)

Episodic memory was also assessed using the Spanish version of the MBT, which employs an associative learning procedure.^41^ Participants learned two lists of 16 semantically paired words under controlled conditions, followed by recall of each list by semantic category and free recall of all 32 items. Delayed recall (free and facilitated) was tested after 30 minutes. Variables of interest included total immediate paired recall (0–32), total immediate free recall (0–32), total delayed paired recall (0–32), and total delayed free recall (0–32).

##### Wechsler Memory Scale–IV (WMS-IV) Logical Memory (LM) Subtest

Narrative episodic memory was assessed using the Spanish version of the LM subtest of the WMS-IV.^42^ Participants were asked to recall two orally presented stories (B and C) immediately after presentation and again after a 20–30 minutes delay, followed by a recognition task. Variables of interest included immediate recall (0–50), delayed recall (0–50), and recognition (0–30).

##### Modified Preclinical Alzheimer’s Cognitive Composite (mPACC)

The PACC is a validated measure of Aβ-related cognitive decline, pooling 4 z-transformed neuropsychological scores.^43^ In this study, we used the mPACC, which was developed for optimization using a semantic fluency measurement instead of the MMSE, enhancing sensitivity to early cognitive changes.^44^ The Free and Cued Selective Reminding Test, Spanish version, was used to assess verbal episodic memory with a controlled learning procedure, considering the total immediate recall (0–48).^40^ The Logical Memory subtest in the Wechsler Memory Scale-IV, Spanish version, was used to assess narrative episodic memory, considering the total delayed recall (0–50).^42^ The Coding subtest in the Wechsler Adult Intelligence Scale-IV, Spanish version, was used to assess processing speed and attention (0–135).^45^ The Semantic Fluency Test, Spanish version, was used to assess executive control through semantic verbal fluency (number of correct lexical items produced in 1 minute within the category “animals”).^46^

##### Amsterdam Instrumental Activities of Daily Living Questionnaire (A-IADL-Q)

Functional abilities in instrumental activities of daily living were assessed using the Spanish A-IADL-Q, a measure sensitive to early functional changes associated with incipient dementia.^47,48^ For this study, the 30-item short version of the A-IADL-Q was administered to the study partner, providing a brief, yet comprehensive assessment of daily functioning.

##### Clinical Dementia Rating (CDR)

The Spanish version of the CDR was used to assess global cognition and daily functioning across six domains, providing a global score from 0 (no impairment) to 3 (severe dementia).^49^

##### Mini Mental State Examination (MMSE)

Global cognition was screened using the Spanish version of the MMSE, which assesses orientation, registration, attention, recall, language, and constructive praxis (0–30).^50^

##### Subjective Cognitive Decline Questionnaire (SCD-Q)

The Spanish SCD-Q was used to assess and quantify SCD, completed independently by both participant and study partner.^51^ The questionnaire includes three initial yes/no items and 24 questions addressing everyday cognitive difficulties. Categorically defined SCD status was determined based on the first item (“Do you perceive memory or cognitive difficulties?” for participants; “Do you perceive he/she has memory or cognitive difficulties?” for study partners). Total scores, restricted to the memory domain were used to quantify subjective memory decline (0–11).

#### Clinical Staging

Clinical staging of ACD was based on the integration of two complementary components: episodic memory and cognitive awareness. Each component was operationalized using robust AD biomarker-based standardized regression-based (SRB) change indices referenced to the Aβ-negative group and adjusted for demographic factors. **Figure 1** provides an overview of this operational framework, illustrating how both components were combined to define the primary study measure. The precise clinical staging criteria applied are described below. Of note, no participant met criteria for cognitive impairment at either baseline or follow-up, defined as: CDR global score > 0; MMSE score < 27, and the conservative application of the neuropsychological criteria,^52^ operationalized as performance < 1.65 SD below the conventional normative mean on at least two of four main measures from the FCSRT.

**Figure 1.**
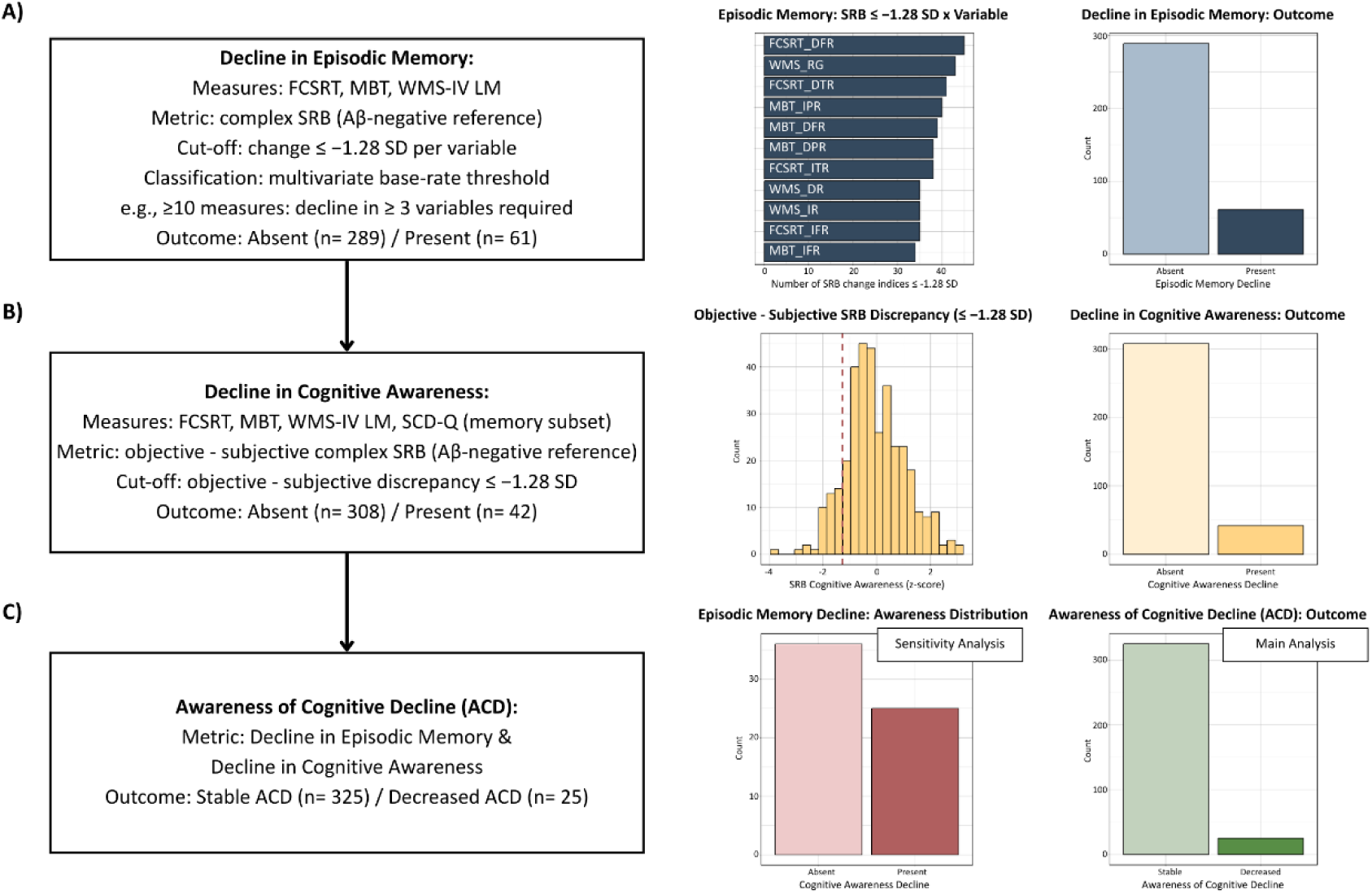
Clinical Staging and Awareness of Cognitive Decline. Flow diagram showing clinical staging and definition of awareness of cognitive decline (ACD). **A)** Episodic memory decline: Participants showed significant longitudinal decline across multiple memory measures (FCSRT, MBT, WMS-IV LM), quantified using AD biomarker–based complex SRB change indices ≤ −1.28 SD (≤10^th^ percentile), adjusted for age, sex, education, and follow-up interval. A multivariate base rate threshold was applied: e.g., participants with ≥10 measures required ≥3 below cut-off. The left bar plot shows the number of SRB change indices below the cut-off for each variable. The right bar plot shows the final classification. **B)** Decline in cognitive awareness: Defined as a discrepancy between objective and subjective cognition (objective–subjective SRB change ≤ −1.28 SD). Objective cognition was a composite of episodic memory SRB indices; subjective cognition was derived from the memory subset of the SCD-Q (inverted to align direction). Lower discrepancy values indicated overestimation of performance (anosognosia). The left histogram shows the distribution of the objective-subjective discrepancy along the cut-off used. The right bar plot shows the final classification. **C)** ACD: Decreased ACD was operationalized as the co-occurrence of (i) episodic memory decline and (ii) decline in cognitive awareness. The left bar plot shows the distribution of decline in cognitive awareness within the subset of participants with episodic memory decline; this classification was evaluated in sensitivity analysis. The right bar plot shows the final classification across the full sample; evaluated in the main analysis.

##### Decline in Episodic Memory

Participants were required to meet criteria for significant cognitive decline across multiple episodic memory variables derived from three tests: four main variables from the FCSRT, four main variables from the MBT, and three from the WMS-IV LM. Decline was operationalized using robust AD biomarker-based complex standardized regression-based (SRB) change indices, defined as a change ≤ −1.28 SD (≤ 10^th^ percentile) per variable. Complex SRB change indices were determined following previously validated methods for developing robust longitudinal neuropsychological references within the ALFA+ cohort.^36^ Briefly, SRB indices were referenced to the Aβ-negative group and adjusted for age, sex, and education, and longitudinal time interval (years). Following prior ALFA+ work, a multivariate base-rate threshold was applied to categorically define episodic memory decline.^53^ Participants with ≥10 available variables were required to show decline on at least three variables, whereas those with <10 variables available were required to show decline on at least two variables, with all 350 participants having complete longitudinal data for the four main FCSRT variables.

##### Decline in Cognitive Awareness

Participants were required to meet the following criterion: significant decline in cognitive awareness, operationalized as a longitudinal discrepancy between objective and subjective cognition, defined using an AD biomarker-based complex SRB change ≤ −1.28 SD (≤ 10^th^ percentile). Objective cognition was defined using a composite of multiple episodic memory robust complex SRB longitudinal indices (FCSRT, MBT, and WMS-IV LM). Subjective cognition was defined using a robust complex SRB longitudinal index of subjective memory decline reports (SCD-Q, memory domain), also referenced to the Aβ-negative group and adjusted for age, sex, education, and longitudinal time interval (years). Resulting SRB subjective scores were inverted (multiplied by −1) to align with the direction of objective cognition, such that higher transformed values represent better self-perceived cognitive performance. The discrepancy was then computed as objective minus subjective SRB change indices, whereby values close to 0 indicate accurate awareness of longitudinal cognitive performance, higher values indicate underestimation of cognitive performance (i.e., hypernosognosia direction: greater subjective complaint than objective decline), and lower values indicate overestimation of cognitive performance (i.e., anosognosia direction: more objective decline than subjectively perceived).

##### Awareness of Cognitive Decline (ACD)

Participants who concurrently met both criteria, (i) decline in episodic memory and (ii) decline in cognitive awareness, were classified as “decreased ACD”, whereas those who did not meet both criteria were classified as “stable ACD”. Of note, the stable ACD classification does not imply the absence of episodic memory decline or the absence of decline in cognitive awareness in absolute terms. Rather, it reflects the failure to concurrently meet both operationalized criteria within the defined staging framework (i.e., significant episodic memory decline and significant decline in cognitive awareness). Participants classified as stable ACD may include individuals with decline in one, but not both components, as well as those showing no decline in either domain. This label should therefore be interpreted as the absence of concurrent decline across both dimensions.

### Statistical Analysis

Analyses were conducted using R 4.2.1 (RStudio 2022.07.1) with nominal p-values < 0.05 considered statistically significant, except for neuroimaging analyses which used SPM 12 (MATLAB 2022b), where results were considered significant at nominal p-values < 0.005 with a cluster-extent threshold > 100 voxels.

### Characteristics of the Participants

Baseline characteristics of the two groups (stable ACD and decreased ACD; Figure 1, panel C) were compared using univariate ANOVAs for continuous variables and Chi-square tests for categorical variables.

### Robust Normative Longitudinal Neuropsychological Change

Neuropsychological variables used for clinical staging were summarized descriptively and compared between groups using t-tests, with effect sizes quantified by Cohen’s d. Associations of ACD with clinical measurements were evaluated using linear regression models.

### Associations with Clinical Measurements

Clinical measures included mPACC, study partner reported subjective memory decline (SCD-Q), and study partner reported functional performance (A-IADL-Q). mPACC, SCD-Q, and A-IADL-Q were defined using AD biomarker-based robust longitudinal complex SRB indices adjusted for age, sex, education, and time. Clinical measures were entered as outcomes in independent models and ACD classification as the main predictor. Of note, the mPACC composite includes two neuropsychological measures (FCSRT total immediate recall and WMS-IV LM total delayed recall) that are also components of the episodic memory criteria used for ACD classification; therefore, primary mPACC analyses should be interpreted in the context of this measurement overlap.

### Associations with Core AD Biomarkers

Associations of ACD with fluid biomarkers (plasma and CSF) were evaluated using linear regression models. Plasma and CSF biomarkers used for statistical analyses were acquired at baseline visit. Log10-transformed fluid biomarkers were entered as outcomes in independent models and ACD classification as the main predictor, adjusting for age and sex. Results were reported including standardized β coefficients with 95% confidence intervals, *p*-values, nominal and adjusted R², and the Akaike Information Criterion (AIC).

Associations of ACD with neuroimaging biomarkers were evaluated using voxel-wise linear regression models. Aβ and tau PET scans used for voxel-wise statistical analyses were acquired at follow-up visit. PET neuroimaging data were entered as outcomes in independent models, with ACD classification as the main predictor, adjusting for age and sex. Voxel-wise neuroimaging models employed an explicit grey matter mask, further excluding the cerebellum. Statistical results were reported at both the cluster and peak levels, including cluster size (k), *p*-value, *T*-value, and peak MNI coordinates (x, y, z), alongside anatomical labels at a representative cluster-level, using 1 mm³ resolution labeling from the Automated Anatomical Labelling Atlas 3.

### Sensitivity Analysis

Sensitivity analyses were restricted to participants with episodic memory decline. Associations between ACD classification and clinical and multimodal AD biomarkers were evaluated using the same methods described above, with outcomes modeled without adjustment for age or sex, as between-group differences on these variables were not observed when restricting the sample to participants with episodic memory decline. These analyses aimed to determine whether, at the same threshold of objective episodic memory decline, individuals exhibiting decreased ACD differed from those maintaining stable ACD. Sensitivity analyses evaluating mPACC attenuated possible confounding driven by measurement overlap, as both ACD groups exhibited episodic memory decline according to the clinical staging criteria, which included the neuropsychological measures shared with the mPACC (FCSRT total immediate recall and WMS-IV LM total delayed recall). However, given that ACD classification inherently incorporates these shared measures through the objective-subjective discrepancy index, mPACC findings should be interpreted in the context of an attenuated, yet persistent measurement dependency. The primary reason for evaluating mPACC, despite its dependency with the clinical staging criteria, is that both PACC and mPACC versions have been established as primary endpoints in AD clinical trials, making them clinically relevant measures with sensitivity to underlying Aβ pathology for detecting preclinical cognitive changes.

### Data Availability

The data supporting the findings of this study are available from the corresponding authors, D.L.-M., G.S.-B., or O.G.-R., on a reasonable request.

## Results

### Characteristics of the Participants

A total of 350 CU middle-aged participants enrolled in the ALFA+ cohort were included in the present study, with a mean longitudinal time interval of 3.30 years (standard deviation = 0.52). According to clinical staging criteria, 325 individuals (93%) were classified as exhibiting stable ACD, and 25 (7%) as exhibiting decreased ACD over the follow-up period. Baseline characteristics are presented in **Table 1**. Significant baseline differences were observed between ACD groups. The group exhibiting decreased ACD was slightly older (*p*-value = 0.006), demonstrated a greater prevalence of CSF Aβ positivity (p-value = 0.012), higher plasma p-tau217 concentration (*p*-value = 0.011), higher CSF p-tau181/Aβ42 ratio (*p*-value < 0.001), higher Aβ PET load (*p*-value < 0.001), and lower performance on episodic memory (*p*-value = 0.006). No significant baseline differences were found for sex, education, *APOE-ε4* status, MMSE, and subjective memory decline total score. Within the stable ACD group, categorically defined self-reported SCD was reported by 89 of 325 individuals (27%) at baseline and increased to 108 of 325 (33%) at follow-up. Within the decreased ACD group, self-reported SCD was present in 9 of 25 individuals (36%) at baseline, a prevalence that remained unchanged at follow-up. No significant group differences for categorically defined self-reported SCD were observed at baseline or follow-up.

**Table 1.** Baseline Characteristics of the Participants.

|  | n | Full Sample<br>(n= 350) | Stable ACD<br>(n= 325) | Decreased ACD<br>(n= 25) | p-value |
| --- | --- | --- | --- | --- | --- |
| *Demographic* |  |  |  |  |  |
| Age, mean (SD) | 350 (325 / 25) | 60.93±4.72 | 60.74±4.63 | 63.41±5.30 | **0.006** |
| Sex [female], n (%) | 350 (325 / 25) | 211 (60.29) | 194 (59.69) | 17 (68) | 0.545 |
| Education (years), mean (SD) | 350 (325 / 25) | 13.53±3.48 | 13.59±3.48 | 12.64±3.51 | 0.188 |
| *Genetic* |  |  |  |  |  |
| *APOE-ε4* [carriership], n (%) | 350 (325 / 25) | 191 (54.57) | 179 (55.08) | 12 (48) | 0.634 |
| *Biomarker* |  |  |  |  |  |
| Plasma p-tau217 (pg/mL), mean (SD) | 349 (324 / 25) | 0.1530±0.0899 | 0.1496±0.0885 | 0.1971±0.0975 | **0.011** |
| CSF p-tau181/Aβ42 (hybrid ratio), mean (SD) | 350 (325 / 25) | 0.0144±0.0109 | 0.0138±0.0100 | 0.0229±0.0173 | **< 0.001** |
| CSF Aβ-positive (Aβ42/Aβ40), n (%) | 350 (325 / 25) | 122 (34.86) | 107 (32.92) | 15 (60) | **0.012** |
| [<sup>18</sup>F]flutemetamol PET (Centiloids), mean (SD) | 304 (284 / 20) | 3.5234±17.2493 | 2.2762±15.7173 | 21.2346±26.6833 | **< 0.001** |
| *Clinical* |  |  |  |  |  |
| mPACC, mean (SD) | 350 (325 / 25) | -0.01±0.67 | 0.04±0.65 | -0.64±0.70 | **< 0.001** |
| MMSE, mean (SD) | 350 (325 / 25) | 29.18±0.92 | 29.20±0.90 | 28.88±1.09 | 0.094 |
| Episodic Memory Composite, mean (SD) | 350 (325 / 25) | -0.01±0.71 | 0.02±0.70 | -0.38±0.77 | **0.006** |
| SCD-Q (Memory), mean (SD) | 350 (325 / 25) | 1.55±2.13 | 1.56±2.16 | 1.40±1.76 | 0.713 |
| Study Partner SCD-Q (Memory), mean (SD) | 349 (324 / 25) | 0.68±1.25 | 0.64±1.22 | 1.12±1.59 | 0.065 |
| Study Partner A-IADL-Q, mean (SD) | 350 (325 / 25) | 68.54±1.53 | 68.59±1.50 | 67.89±1.79 | **0.026** |

### Robust Normative Longitudinal Neuropsychological Change

Results of the neuropsychological variables used for clinical staging, stratified by ACD groups (Figure 1, panel C) are presented in **Table 2**. Focusing on the measurements used to define decline in episodic memory. In the full sample, the prevalence of decline per variable (≤ 10th percentile) ranged approximately from 10% to 13% across FCSRT, MBT, and WMS measures, with the highest prevalence observed for FCSRT Total Free Delayed Recall (12.86%), WMS LM: Total Recognition (12.8%) and MBT Total Immediate Paired Recall (11.87%). The group exhibiting decreased ACD showed lower SRB change scores than the stable ACD group across all episodic memory measures examined. The largest difference was observed for MBT Total Immediate Free Recall (*p*-value < 0.001, Cohen’s d = 1.606), followed by FCSRT Total Free Delayed Recall (*p*-value < 0.001, Cohen’s d = 1.464), and WMS LM Total Immediate Recall (*p*-value < 0.001, Cohen’s d = 1.343). Regarding the measures used to define decline in cognitive awareness, the episodic memory composite showed substantially lower SRB change in the decreased ACD group compared with the stable group (*p*-value < 0.001, Cohen’s d = 2.667). Self-reported subjective memory decline showed non-significant group differences. Finally, the cognitive awareness SRB change differed markedly between ACD groups (*p*-value < 0.001, Cohen’s d = 1.945).

**Table 2.** Robust AD Biomarker-based Complex SRB Change Indices.

| Variables | Full Sample (n = 350) |  |  | Stable ACD<br>(N = 315) | Decreased<br>ACD (n = 25) | Group Comparison |  |  |
| --- | --- | --- | --- | --- | --- | --- | --- | --- |
| | n | SRB ≤<br>percentile<br>10<sup>th</sup>, n (%) | SRB, mean<br>(SD) | SRB, mean<br>(SD) | SRB, mean<br>(SD) | t-statistic | p-value | Cohen's d |
| *Measurements used to define decline in episodic memory* |  |  |  |  |  |  |  |  |
| FCSRT: Total Free Immediate Recall | 350 | 35 (10) | -0.072 (1.038) | 0.014 (0.977) | -1.188 (1.182) | 4.958 | **<0.001** | 1.211 |
| FCSRT: Total Immediate Recall | 350 | 38 (10.86) | -0.082 (1.124) | 0.019 (0.956) | -1.399 (2.019) | 3.482 | **0.002** | 1.332 |
| FCSRT: Total Free Delayed Recall | 350 | 45 (12.86) | -0.112 (1.079) | -0.006 (0.986) | -1.485 (1.3) | 5.569 | **<0.001** | 1.464 |
| FCSRT: Total Delayed Recall | 350 | 41 (11.71) | -0.076 (1.197) | 0.041 (0.961) | -1.595 (2.4) | 3.389 | **0.002** | 1.459 |
| MBT: Total Immediate Paired Recall | 337 | 40 (11.87) | -0.043 (1.121) | 0.079 (0.96) | -1.638 (1.73) | 4.806 | **<0.001** | 1.665 |
| MBT: Total Immediate Free Recall | 336 | 34 (10.12) | -0.06 (1.062) | 0.048 (0.979) | -1.534 (1.066) | 6.906 | **<0.001** | 1.606 |
| MBT: Total Delayed Paired Recall | 337 | 38 (11.28) | -0.032 (1.167) | 0.092 (0.998) | -1.653 (1.847) | 4.577 | **<0.001** | 1.618 |
| MBT: Total Delayed Free Recall | 334 | 39 (11.68) | -0.09 (1.089) | -0.003 (1.023) | -1.323 (1.264) | 4.789 | **<0.001** | 1.269 |
| WMS LM: Total Immediate Recall | 336 | 35 (10.42) | -0.129 (1.07) | -0.032 (0.989) | -1.392 (1.291) | 5.049 | **<0.001** | 1.343 |
| WMS LM: Total Delayed Recall | 335 | 35 (10.45) | -0.1 (1.022) | -0.019 (0.957) | -1.143 (1.258) | 4.284 | **<0.001** | 1.146 |
| WMS LM: Total Recognition | 336 | 43 (12.8) | -0.098 (1.03) | -0.012 (0.971) | -1.22 (1.142) | 5.043 | **<0.001** | 1.228 |
| *Measurements used to define decline in cognitive awareness* |  |  |  |  |  |  |  |  |
| Episodic Memory Composite | 350 | 11 (3.14) | -0.087 (0.711) | 0.025 (0.504) | -1.539 (1.245) | 6.239 | **<0.001** | 2.667 |
| Subjective Memory Decline | 350 | 14 (4) | 0.031 (0.984) | 0.057 (0.981) | -0.301 (0.987) | 1.747 | 0.092 | 0.365 |
| Cognitive Awareness | 350 | 42 (12) | -0.056 (1.104) | 0.082 (1.013) | -1.84 (0.533) | 15.934 | **<0.001** | 1.945 |

The table presents AD biomarker-based robust complex SRB longitudinal change indices for the neuropsychological battery, including measurements used to define decline in episodic memory and cognitive awareness. Development of these indices followed methods previously described (see López-Martos, D., et al., 2025). Robust longitudinal complex SRB indices reflect intra-individual cognitive change referenced to the Aβ-negative sample (z-scores) adjusted for age, sex, education, longitudinal time interval. The episodic memory composite was defined using all available SRB indices for each participant to characterize decline in episodic memory. SCD memory represents quantification of cognitive complaints restricted to the memory domain (SCD-Questionnaire), with higher scores indicating greater subjective memory decline. Cognitive awareness was defined as the standardized discrepancy between objective and subjective memory performance, using inverted SCD scores ( $-1 \times \text{SCD}$ ), with lower scores indicating lower awareness. For each variable, the table reports the number of observations (n), not available data (NA), the number of observations and percentage (%) of individuals below the 10<sup>th</sup> percentile ( $\leq -1.28$ SRB), and the mean (SD) values for the full sample. Results are also stratified by ACD groups, with mean (SD), t-statistic, p-value, and Cohen's d for group comparisons. Bold text indicates significant results (p-values $< 0.05$ ).

### Associations with Clinical Measurements

Associations of ACD with clinical measurements are presented in **Table 3**. Decreased ACD was associated with more adverse clinical outcomes, compared to stable ACD. Participants with decreased ACD showed significantly greater longitudinal decline in cognition, as assessed by mPACC (Std. β = -1.421, 95% CI = [-1.801, -1.040], *p*-value < 0.001), greater longitudinal increase in study partner reported subjective memory decline, as assessed by SCD-Q (Std. β = 0.557, 95% CI= [0.152, 0.961], *p*-value = 0.007), and greater longitudinal decline in study partner reported functional performance, as assessed by A-IADL-Q (Std. β = -0.893, 95% CI = [-1.291, -0.495], *p*-value < 0.001).

**Table 3.** Associations of Decreased Awareness of Cognitive Decline with Clinical Measurements.

| Outcomes | n | Std. $\beta$ (95% CI) | $p$-value | R<sup>2</sup> | AIC |
| --- | --- | --- | --- | --- | --- |
| mPACC | 350 (325 / 25) | -1.421 (-1.801, -1.040) | **< 0.001** | 0.134 / 0.132 | 596.35 |
| Study Partner SCD-Q (Memory) | 348 (323 / 25) | 0.557 (0.152, 0.961) | **0.007** | 0.021 / 0.018 | 1044.348 |
| Study Partner A-IADL-Q | 348 (323 / 25) | -0.893 (-1.291, -0.495) | **< 0.001** | 0.053 / 0.051 | 1038.472 |

Results from linear regression models indicating outcomes, number of observations (n; Stable ACD / Decreased ACD), standardized (Std.) $\beta$ coefficients with 95% Confidence Intervals (CI), nominal $p$ -values, R<sup>2</sup> (i.e., nominal/adjusted proportion of explained variance), and Akaike Information Criteria (AIC) measurements. Clinical measures, mPACC, SCD-Q, and A-IADL-Q were determined using longitudinal complex SRB methods adjusted for age, sex, education, and time, entered as outcomes in independent models using ACD classification (0: Stable ACD, 1: Decreased ACD) as the main predictor. Bold text indicates a significant association ( $p$ -value < 0.05).

### Associations with Core AD Biomarkers

#### Plasma and CSF Biomarkers

Associations of ACD with plasma and CSF biomarkers are presented in **Table 4** and **Figure 2**. Compared to the stable ACD group, decreased ACD was associated with greater AD pathological burden, as measured by fluid biomarkers. Among plasma measures, the strongest association was observed for plasma p-tau181 (Std. β = 0.625, 95% CI = [0.217, 1.033], *p*-value = 0.003), followed by plasma p-tau181/Aβ42 ratio (Std. β = 0.612, 95% CI = [0.199, 1.024], *p*-value = 0.004) and plasma p-tau217 (Std. β = 0.582, 95% CI = [0.173, 0.990], *p*-value = 0.005). Among CSF measures, the strongest association was observed for CSF p-tau217 (Std. β = 0.791, 95% CI = [0.402, 1.180], *p*-value < 0.001), closely followed by the CSF p-tau181/Aβ42 ratio (Std. β = 0.721, 95% CI = [0.334, 1.108], *p*-value < 0.001). A smaller but significant association was also observed for CSF p-tau181 (Std. β = 0.420, 95% CI = [0.022, 0.817], *p*-value = 0.038).

**Table 4.**
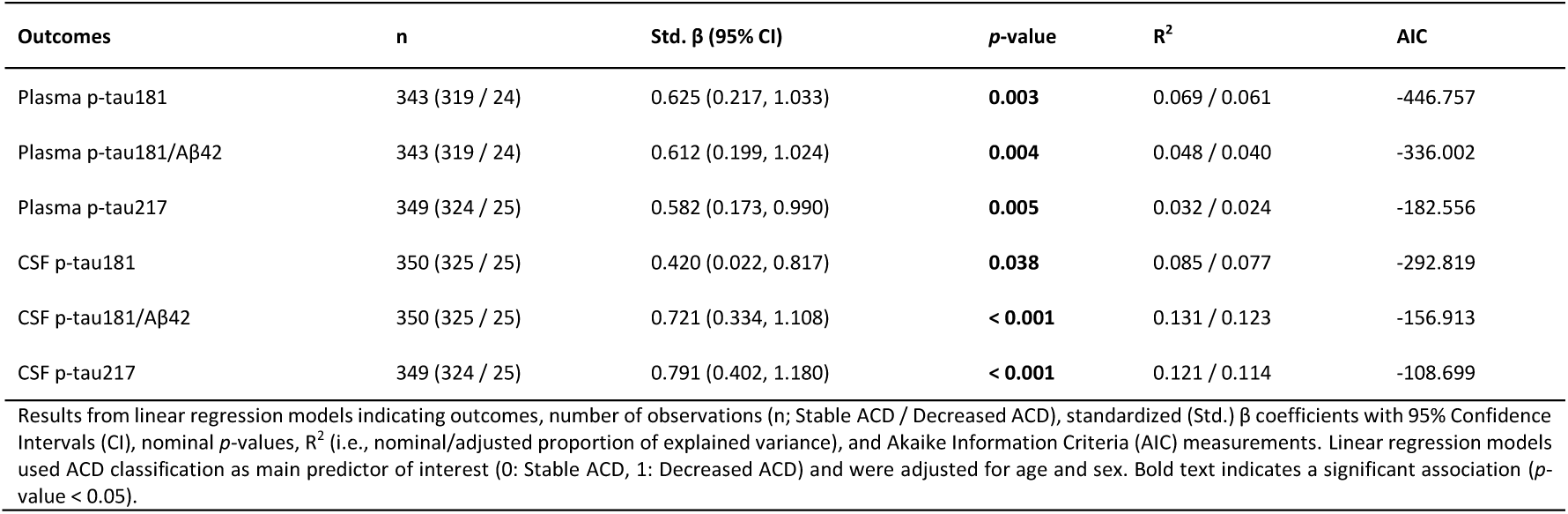
Associations of Decreased Awareness of Cognitive Decline with Plasma and CSF Biomarkers.

**Figure 2.**
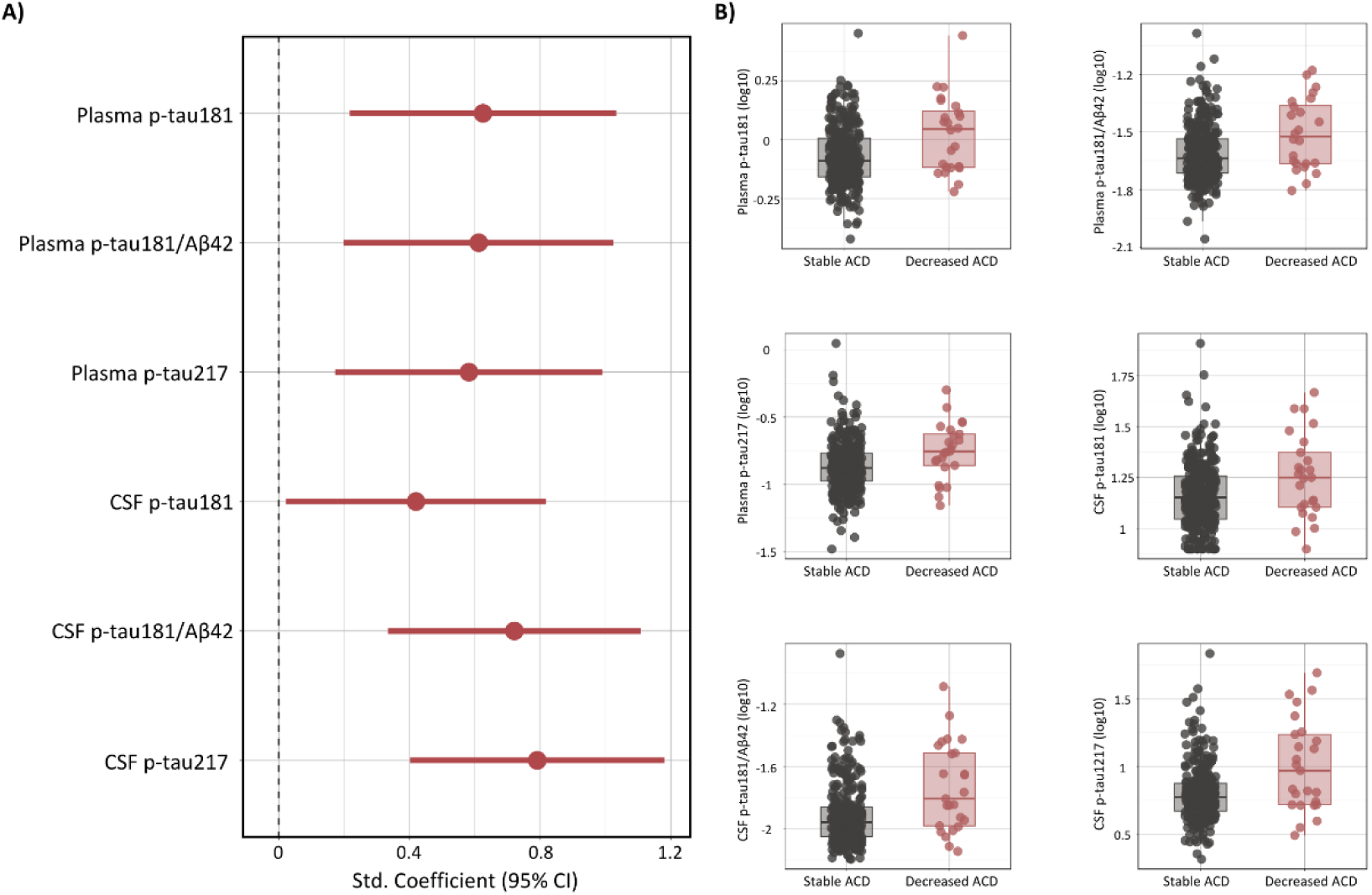
Associations of Decreased Awareness of Cognitive Decline with Plasma and CSF Biomarkers. A) Forest plot showing standardized regression coefficients and 95% confidence intervals for the effect of group (ACD classification; 0: Stable ACD, 1: Decreased ACD) on plasma and CSF biomarker outcomes from linear regression models adjusted for age and sex. B) Raw boxplots displaying the distribution of plasma and CSF biomarkers between groups.

#### PET Biomarkers

Associations of ACD with PET biomarkers are presented in **Table 5** and **Figure 3**. Compared to the stable ACD group, decreased ACD was associated with greater AD pathological burden, as measured by neuroimaging biomarkers. For Aβ PET, the group exhibiting decreased ACD showed higher Aβ burden, primarily in frontal and parietal cortical regions. Additional deposition was observed in lateral temporal areas, reflecting a global, widespread cortical Aβ pattern. For tau PET, the group exhibiting decreased ACD showed higher tau burden largely localized to temporal regions. Within temporal areas, the medial temporal lobe (MTL) exhibited the strongest involvement, with additional contributions from inferior and middle temporal regions. Tau PET deposition was also observed in insular and frontal areas, altogether demonstrating a topographical pattern extending beyond the MTL into neocortical regions.

**Table 5.** Associations of Decreased Awareness of Cognitive Decline with PET Biomarkers.

| Outcomes | n | Anatomical locations | Cluster-level |  | Peak-level |  | Peak MNI |  |  |
| --- | --- | --- | --- | --- | --- | --- | --- | --- | --- |
|  |  |  | k | p-value | T | p-value | x | y | z |
| Aβ PET | 195 (182 / 13) | Middle Frontal Left | 44259 | < 0.001 | 6.115 | < **0.001** | -26 | 8 | 0 |
|  |  | Superior Frontal Right | 191 | 0.109 | 4.416 | < **0.001** | 28 | 26 | 54 |
|  |  | Inferior Temporal Left | 706 | 0.005 | 4.115 | < **0.001** | -48 | -52 | -16 |
|  |  | Thalamus Right | 130 | 0.180 | 4.077 | < **0.001** | 12 | -26 | 6 |
|  |  | Fusiform Right | 105 | 0.226 | 3.851 | < **0.001** | 34 | -4 | -40 |
|  |  | Parahippocampal Left | 185 | 0.114 | 3.588 | < **0.001** | -18 | -38 | -12 |
|  |  | Middle Occipital Left | 156 | 0.144 | 3.520 | < **0.001** | -46 | -72 | 14 |
|  |  | Parahippocampal Right | 173 | 0.125 | 3.477 | < **0.001** | 30 | -36 | -14 |
|  |  | Superior Parietal Right | 114 | 0.208 | 3.128 | **0.001** | 24 | -56 | 66 |
| TAU PET | 87 (79 / 8) | Medial Temporal Lobe Left | 5096 | < 0.001 | 5.740 | < **0.001** | -20 | -8 | -28 |
|  |  | Medial Temporal Lobe Right | 4247 | < 0.001 | 5.667 | < **0.001** | 20 | -4 | -28 |
|  |  | Middle Temporal Right | 774 | 0.044 | 4.337 | < **0.001** | 52 | -66 | 16 |
|  |  | Superior Frontal Left | 317 | 0.179 | 3.640 | < **0.001** | -6 | 26 | 52 |

Results from whole-brain voxel-wise linear regression models indicating outcomes, number of observations (n; Stable ACD / Decreased ACD), cluster-level anatomical location defined using 1mm3 resolution cluster labelling from the Automated Anatomical Labelling Atlas 3, cluster-level voxel size (k), cluster p-values, peak-level T-statistics, peak-level p-values, and peak-level coordinates according to the Montreal Neurological Institute (MNI) space. The voxel-wise neuroimaging models used ACD classification as main predictor of interest (0: Stable ACD, 1: Decreased ACD) and were adjusted for age and sex. Bold text indicates a significant association (nominal p-values < 0.005 at the peak-level with a cluster-level threshold correction > 100 voxels).

**Figure 3.**
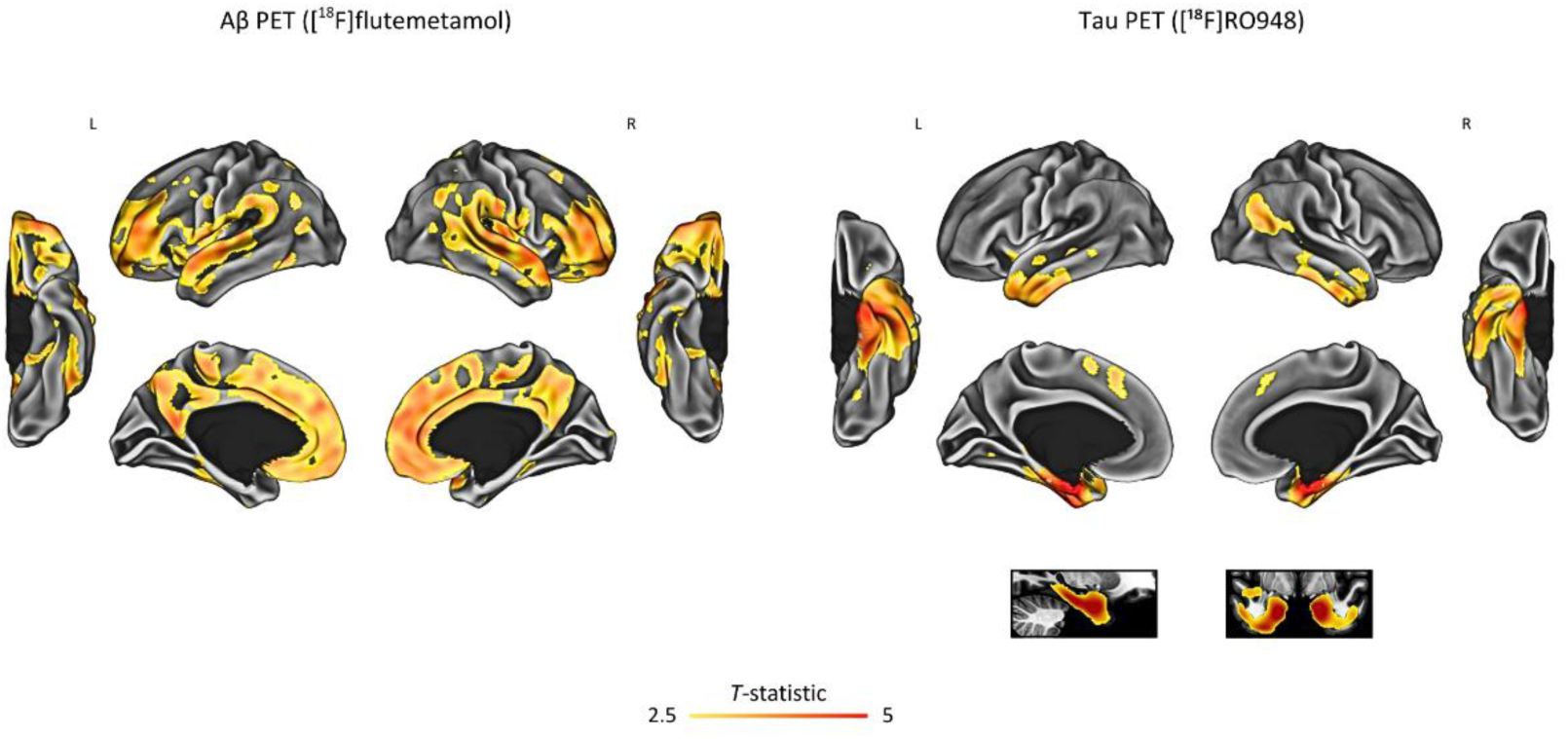
Associations of Decreased Awareness of Cognitive Decline with PET Biomarkers. Results from voxel-wise linear regression models showing the effect of group (ACD classification; 0: Stable ACD, 1: Decreased ACD) on PET biomarker outcomes. The figure displays cortical surface-based projections following anatomical convention, where the left side of the image corresponds to the left hemisphere of the brain. Additionally, voxel-wise results are shown on T1-weighted anatomical sections. Color bars indicate *T*-statistics from vertex-wise projection into the cortical surface, with results shown at nominal *p*-values < 0.005 accounting for a cluster-size threshold of k > 100 voxels. Models were adjusted for age and sex.

### Sensitivity Analysis

#### Characteristics of the Participants

Sensitivity analysis was restricted to participants with episodic memory decline, regardless of cognitive awareness status. Specifically, 61 participants (17%) were classified as exhibiting episodic memory decline (Figure 1, panel A), of whom 25 (41%) also showed decline in cognitive awareness, meeting criteria for decreased ACD (Figure 1, panel C). Baseline characteristics are provided in Supplementary Table 1. Compared to the group with episodic memory decline but stable ACD, the group with decreased ACD showed higher Aβ PET burden (*p*-value = 0.035) and lower mPACC (*p*-value = 0.007). No significant baseline differences were found for age, sex, education, *APOE*-ε4, plasma p-tau217, CSF p-tau181/Aβ42, CSF Aβ-positive (Aβ42/Aβ40), MMSE, episodic memory composite, SCD-Q (memory), Study Partner SCD-Q (Memory), and Study Partner A-IADL-Q.

#### Associations with Clinical Measurements

Associations of ACD with clinical measurements are presented in Supplementary Table 2. Compared to the group with episodic memory decline but stable ACD, the group with decreased ACD showed significantly greater longitudinal decline in cognition, as assessed by mPACC (Std. β = -0.591, 95% CI = [-1.093, -0.088], *p*-value = 0.022). No significant differences were observed for study partner reports, considering both SCD-Q and A-IADL-Q.

#### Associations with Core AD Biomarkers

#### Plasma and CSF Biomarkers

Associations of ACD with plasma and CSF biomarkers are presented in Supplementary Table 3 and Supplementary Figure 1. Compared to the group with episodic memory decline but stable ACD, the group with decreased ACD showed higher CSF p-tau217 concentration (Std. β = 0.564, 95% CI = [0.059, 1.068], *p*-value = 0.029) and higher plasma p-tau181/Aβ42 ratio (Std. β = 0.520, 95% CI = [0.006, 1.034], *p*-value = 0.047). No significant differences were observed for plasma p-tau181, plasma p-tau217, CSF p-tau181, and CSF p-tau181/Aβ42 ratio.

#### PET Biomarkers

Associations of ACD with PET AD biomarkers are presented in Supplementary Table 4 and Supplementary Figure 2. Compared to the group with episodic memory decline but stable ACD, the group with decreased ACD showed greater Aβ PET burden, primarily in frontoparietal and cortical temporal regions, alongside greater tau PET burden, primarily in MTL and extending into inferior and middle temporal regions, with additional localized deposition in insular cortex.

## Discussion

The present study provided a comprehensive clinical and biological characterization of decreased ACD in CU individuals across aging and the preclinical stage of AD. We integrated longitudinal objective neuropsychological performance with subjective reports of cognitive decline to evaluate changes in self-awareness in relationship to AD pathology. Specifically, we operationalized decreased ACD as the co-occurrence of decline in episodic memory and cognitive awareness. Our findings revealed that decreased ACD captures a clinically and biologically meaningful phenotype associated with steeper cognitive and functional decline, as well as greater AD multimodal biomarker levels. Notably, among individuals with episodic memory decline, those additionally exhibiting decreased ACD demonstrated steeper cognitive decline and greater AD pathological burden, underscoring that self-awareness carries relevant clinical value beyond common memory performance thresholds. Together, these findings support that AD-related subtle alterations in awareness emerge alongside cognitive decline, highlighting the relevance of assessing cognitive awareness to enhance early detection and risk stratification in aging and preclinical AD.

Individuals exhibiting decreased ACD showed greater longitudinal decline in global cognitive performance, as measured by the mPACC, a composite of episodic memory and executive function sensitive to early Aβ deposition, widely used to capture AD-related decline, and also considered as a primary endpoint in AD clinical trials.^54^ This finding highlights an early dissociation between objective decline and subjective perception that mirrors the course of self-awareness in AD progression, yet already detectable in aging and preclinical AD.^14^ Consistently, study partner reports revealed significant associations of decreased ACD with more detrimental cognitive and functional changes. Study partner reports of memory decline, assessed via SCD-Q, increased more steeply over time in the group exhibiting decreased ACD, emphasizing a key the distinction between participant and partner reports, critical for early detection, monitoring, and risk stratification.^55^ Study partner reports of everyday functioning, measured with A-IADL-Q, declined more over time in the group exhibiting decreased ACD, suggesting that changes in cognitive awareness may track with subtle changes in functional capacity prior to clinical onset.^56^ Together, these results indicate that decreased ACD reflects greater cognitive and functional vulnerability, supporting its clinical relevance for identifying CU individuals who might benefit most from early monitoring and intervention despite minimal subjective concern.

Fluid biomarkers revealed a greater overall burden of AD pathology associated with decreased ACD, reflecting early pathophysiological processes. Specifically, the pattern of associations reflected Aβ aggregation and tau phosphorylation, key biological processes in preclinical AD. ^57–61^ This biomarker profile suggests that decreased ACD reflects a more advanced stage of underlying AD progression. Fluid biomarkers, which become abnormal earlier than PET imaging, begin to change and become detectable across the early AD *continuum*, indicating an elevated risk of clinical and biological disease progression.^62–64^ We observed relatively stronger associations for CSF p-tau217 and CSF p-tau181/Aβ42 ratio. Specifically, the prominence of CSF p-tau217 may reflect more advanced pathological progression, reflecting both Aβ and tau PET deposition more sensitively than CSF p-tau181.^65^ The strength of the CSF p-tau181/Aβ42 hybrid ratio suggests that the synergistic expression of both pathologies, rather than phosphorylated tau alone, maximizes the detection of key pathological changes linked to both Aβ and tau PET, ^66,67^ enhancing characterization of early pathological progression in decreased ACD. Therefore, these findings suggest that concurrent decline in memory and awareness is most closely linked to fluid AD biomarkers reflecting more advanced disease progression.^63^ The relatively stronger associations observed for CSF compared with plasma measures likely reflect CSF’s higher signal-to-noise ratio and greater sensitivity in distinguishing clinically meaningful phenotypes, whereas plasma biomarkers may show broader fluctuations across the entire AD-enriched CU cohort. Nevertheless, differences in association strength should be interpreted cautiously, as they reflect the distinct temporal dynamics of AD biomarkers and specific assay characteristics.^63^ The significant associations observed for plasma biomarkers underscore their potential as scalable, minimally invasive tools for supporting biologically defined AD and more comprehensive characterization of early clinical phenotypes at-risk of progression,^68–71^ with implications for both research and clinical practice. ^59,72,73^

Neuroimaging biomarkers provided further key insights. Individuals exhibiting decreased ACD showed higher global Aβ PET burden, with prominent involvement of Aβ in frontoparietal cortices and additional deposition in temporal regions, consistent with early-to-intermediate stages of cortical Aβ accumulation.^74–78^ This group also showed increased tau PET deposition, primarily localized to the MTL, with notable extension into inferior and middle temporal cortices, and additional insular and frontal involvement. Topographically, this pattern of tau PET is consistent with brain regions known to be affected in early-to-mid stages of tau deposition, with positivity in Braak stages III-IV predominantly observed in Aβ-positive individuals and typically accompanied by MCI.^58,62,74,76,79^ Together, these findings indicate regional patterns of Aβ and tau pathology directly consistent with AD biomarker-staging, supporting decreased ACD as an early clinical feature linked to the AD-related neuropathological change.^80–82^ Specifically, this encompassed Aβ and tau PET deposition in key hubs of the default mode network (DMN), frontoparietal network (FPN), and salience network (SN); including temporal structures predominantly associated with episodic memory, as well as frontal, parietal, and insular cortices, together with midline regions such as the precuneus and cingulate cortex, notably involved in complementary aspects of cognitive control and self-referential processing. Consequently, the presence of greater AD pathology within these key regions suggests a dual disruption of primarily MTL-dependent memory circuits alongside more distributed brain networks supporting meta-cognition, affecting functionally integrated brain regions critical for awareness of episodic memory.^16,28,83–96^

Sensitivity analyses were conducted restricting the sample of participants to those exhibiting episodic memory decline, aiming to isolate the specific contribution of cognitive awareness. These results positioned decreased ACD as a distinctive clinical feature associated with AD-related clinical and biological outcomes. Specifically, compared to individuals with memory decline but stable ACD, those exhibiting decreased ACD showed greater longitudinal decline on the mPACC, revealing increased cognitive vulnerability. Differences in study partner-reported SCD-Q and A-IADL-Q were absent, which is somehow expected, considering that both groups shared comparable levels of objective memory decline; from the study partners perspective, cognitive and functional changes appear similar between groups, since the defining feature of decreased ACD is reduced subjective self-appraisal. Considering biomarkers, CSF p-tau217 showed the strongest association in fluid, supporting findings from the main analysis, yet other plasma and CSF biomarkers showed more limited or nonsignificant associations, likely reflecting the more limited statistical power and narrower clinical contrast addressed with the sensitivity analyses. Similarly, neuroimaging findings revealed a consistent topographical pattern of frontoparietal Aβ PET and temporal tau PET burden, preserving the primary regional distribution observed in the main analyses, though slightly more restricted in spatial extent. Together, these findings show that, among individuals exhibiting subtle episodic memory decline, those with decreased self-awareness represent a distinct profile at higher risk.

These findings have significant implications for early AD detection, contributing to growing evidence showing that subtle alterations in awareness emerge earlier than previously recognized.^8^ Specifically, decreased ACD was associated with greater AD pathology and more adverse longitudinal clinical outcomes, challenging the prevalent assumption that decline in awareness is strictly confined to symptomatic stages.^8^ This observation has important clinical consequences. Failure to explicitly account for decreased ACD in clinical assessment may lead to systematic overlooking of the most vulnerable population; those underreporting cognitive symptoms and delaying seeking medical attention due to AD-related metacognitive decline.^18,19^ Our study suggests that awareness cannot be assumed to be preserved among CU individuals, particularly in those exhibiting subtle cognitive decline. We found that approximately 40% of those with memory decline also exhibited decline in awareness, and this specific distinction reflected steeper cognitive decline and greater AD pathology. Notably, these individuals presented with minimal subjective concern despite pronounced objective memory decline, a clear dissociation that characterizes a distinct, high-risk clinical phenotype. This pattern suggests that subtle anosognosia may emerge as a clinically meaningful signal from the earliest stages of preclinical AD, challenging the conceptualization of SCD as “the early stage” preceding MCI. Integrating objective and subjective cognitive performance into assessment of ACD may provide a crucial extension of current diagnostic frameworks, opening new avenues for enriching prevention trials, implementing more representative prevention strategies, and supporting personalized monitoring approaches.

Several study limitations should be acknowledged. Criteria used for clinical staging resulted, by definition, in a relatively low-to-modest prevalence of episodic memory decline (approximately 17% of the sample), though decline in awareness in this subset had an important prevalence (approximately 40%). This selective focus on the lower end of neuropsychological distributions enhances specificity for detecting early pathological changes but necessarily restricts sample size and may underrepresent milder phenotypes, potentially limiting generalizability of these results.^36^ Additionally, the relatively small sample with available PET imaging further warrants cautious interpretation of our neuroimaging findings, where replication in larger cohorts will be important to confirm these regional patterns. Importantly, longer follow-up periods will be necessary to evaluate the predictive value of ACD within the CU range for progression to MCI and dementia.^14^ Similarly, future work should explore whether subtle decline in awareness is primarily domain-specific, driven by a subclinical amnestic variant of the hippocampal type,^53^ or extends beyond episodic memory to other cognitive domains, further investigating potential relationships with emerging neuropsychiatric symptoms that may co-occur in early AD stages.^97,98^ Finally, our cohort characteristics also require consideration, since the sample was enriched for AD risk factors, and consisted primarily of middle-aged, highly educated CU volunteers from Barcelona. These demographic and genetic characteristics may limit the generalizability of our findings to broader clinical populations and more heterogeneous cohorts, particularly those with different educational and cultural backgrounds, genetic profiles, or geographic origins.

In conclusion, this study integrated longitudinal objective neuropsychological performance with subjective reports of cognitive decline to characterize decreased ACD in CU individuals. By defining decreased ACD as concurrent decline in memory and awareness, we identified a clinically meaningful phenotype consistently associated with greater AD pathology across fluid and neuroimaging biomarkers. Sensitivity analyses confirmed that, at the same threshold of memory decline, decreased ACD reflects greater AD pathology, reflecting a distinctive clinical and biological phenotype. These findings highlight the potential value of integrating assessment of cognitive awareness into clinical frameworks for early detection and risk stratification in preclinical AD.

## Acknowledgments

This publication is part of the ALFA study. The authors would like to express their most sincere gratitude to the ALFA project participants and relatives without whom this research would not have been possible. The authors also thank Roche Diagnostics International Ltd for providing the kits used to measure CSF biomarkers, the laboratory technicians at the Clinical Neurochemistry Laboratory in Mölndal, Sweden, for performing the analyses, Eli Lilly and Company for providing the measurements of the in-house assay for plasma p-tau217, GE Healthcare for supplying the [¹⁸F]flutemetamol doses, and F. Hoffmann-La Roche Ltd for sponsoring the tau PET study.

COBAS and ELECSYS are trademarks of Roche. All other product names and trademarks are the property of their respective owners. The NeuroToolKit is a panel of exploratory prototype assays designed to robustly evaluate biomarkers associated with key pathologic events characteristic of AD and other neurological disorders, used for research purposes only and not approved for clinical use (Roche Diagnostics International Ltd, Rotkreuz, Switzerland). The Elecsys β-Amyloid(1–42) CSF and Elecsys Phospho-Tau (181P) CSF assays are approved for clinical use.

Collaborators of the ALFA study are: Federica Anastasi, Annabella Beteta, Marta del Campo, Lidia Canals, Alba Cañas, Irene Cumplido-Marinero, Carme Deulofeu, Ruth Dominguez, Maria Emilio, Karine Fauria, Ana Fernández-Angulo, Sherezade Fuentes, Marina García, Patricia Genius, Armand González-Escolar, Laura Hernández, Felipe Hernández-Viadel, Gema Huesa, Jordi Huguet, Laura Iglesias, Esther Jiménez, Ferran Lugo, Paula Marne, Tania Menchón, José Luis Molinuevo, Paula Ortiz-Romero, Wiesje Pelkmans, Albina Polo, Sandra Pradas, Blanca Rodríguez-Fernández, Iman Sadeghi, Lluís Solsona, Anna Soteras, Laura Stankeviciute, Núria Tort-Colet, Marc Vilanova, Natalia Vilor-Tejedor.

## Funding

The research leading to these results has received funding from “la Caixa” Foundation under agreement LCF/PR/SC22/68000001, the Alzheimer’s Association, and an international anonymous charity foundation through the TriBEKa Imaging Platform project (TriBEKa-17-519007). RC received funding from the MCIN/AEI/10.13039/501100011033/FEDER, EU, through the project PID2021-125433OA-100 and from the grant RYC2021-031128-I, funded by MCIN/AEI/10.13039/501100011033 and the European Union NextGenerationEU/PRTR; MS-C received funding from the ERC under the EU’s Horizon 2020 research and innovation program (grant no. 948677), ERA PerMed-ERA NET and the Generalitat de Catalunya (Departament de Salut) through project no. SLD077/21/000001, projects PI19/00155 and PI22/00456, funded by Instituto de Salud Carlos III (ISCIII) and co-funded by the EU (FEDER), and from a fellowship from ‘la Caixa’ Foundation (ID 100010434) and the EU’s Horizon 2020 research and innovation program under the Marie Skłodowska-Curie (grant no. 847648 (LCF/BQ/PR21/11840004)); AB-S received funding from the Alzheimer’s Association Clinician Scientist Fellowship (AACSF) Program, through the grant AACSF-23-1145154; HZ is a Wallenberg Scholar and a Distinguished Professor at the Swedish Research Council supported by grants from the Swedish Research Council (#2023-00356, #2022-01018 and #2019-02397), the European Union’s Horizon Europe research and innovation programme under grant agreement No 101053962, Swedish State Support for Clinical Research (#ALFGBG-71320), the Alzheimer Drug Discovery Foundation (ADDF), USA (#201809-2016862), the AD Strategic Fund and the Alzheimer’s Association (#ADSF-21-831376-C, #ADSF-21-831381-C, #ADSF-21-831377-C, and #ADSF-24-1284328-C), the European Partnership on Metrology, co-financed from the European Union’s Horizon Europe Research and Innovation Programme and by the Participating States (NEuroBioStand, #22HLT07), the Bluefield Project, Cure Alzheimer’s Fund, the Olav Thon Foundation, the Erling-Persson Family Foundation, Familjen Rönströms Stiftelse, Familjen Beiglers Stiftelse, Stiftelsen för Gamla Tjänarinnor, Hjärnfonden, Sweden (#FO2022-0270), the European Union’s Horizon 2020 research and innovation programme under the Marie Skłodowska-Curie grant agreement No 860197 (MIRIADE), the European Union Joint Programme – Neurodegenerative Disease Research (JPND2021-00694), the National Institute for Health and Care Research University College London Hospitals Biomedical Research Centre, the UK Dementia Research Institute at UCL (UKDRI-1003), and an anonymous donor; KB is supported by the Swedish Research Council (#2017-00915 and #2022-00732), the Swedish Alzheimer Foundation (#AF-930351, #AF-939721, #AF-968270, and #AF-994551), Hjärnfonden, Sweden (#ALZ2022-0006, #FO2024-0048-TK-130 and FO2024-0048-HK-24), the Swedish state under the agreement between the Swedish government and the County Councils, the ALF-agreement (#ALFGBG-965240 and #ALFGBG-1006418), the European Union Joint Program for Neurodegenerative Disorders (JPND2019-466-236), the Alzheimer’s Association 2021 Zenith Award (ZEN-21-848495), the Alzheimer’s Association 2022-2025 Grant (SG-23-1038904 QC), La Fondation Recherche Alzheimer (FRA), Paris, France, the Kirsten and Freddy Johansen Foundation, Copenhagen, Denmark, Familjen Rönströms Stiftelse, Stockholm, Sweden, and an anonymous philanthropist and donor; JDG was supported by the Spanish Ministry of Science and Innovation (RYC–2013–13054), received research support from the EU/EFPIA Innovative Medicines Initiative Joint Undertaking AMYPAD (grant agreement 115952), EIT Digital (grant 2021), and from Ministerio de Ciencia y Universidades (grant agreement RTI2018–102261); OG-R received support from the grant IJC2020-043417-I, funded by MCIN/AEI/10.13039/501100011033 and the European Union NextGenerationEU/PRTR, and receives funding from Instituto de Salud Carlos III (ISCIII) through the projects “PI19/00117” and “PI24/00116”, co-funded by the European Union/FEDER, and through the project “CB16/10/00417”; GS-B receives support from the grant CP23/00039 (Miguel Servet), funded by the Instituto de Salud Carlos III (ISCIII) and co-funded by the European Union/FSE+, and receives funding from the MCIN/AEI/10.13039/501100011033 through the project PID2020-119556RA-I00 and through the project PID2024-163071OB-I00, funded by MICIU/AEI/10.13039/501100011033 and by the European Union FEDER.

## Disclosure

DL-M, RC, MS, AG-E, AH-B, HP-G, MM-A, AB-S, and CM have nothing to disclose; MS-C has served as a consultant and at advisory boards for Roche Diagnostics International Ltd and Grifols S.L., has given lectures in symposia sponsored by Roche Diagnostics, S.L.U and Roche Farma, S.A., and was granted with a project funded by Roche Diagnostics International Ltd; GS has received speaker fees from Springer, GE Healthcare, Biogen, Esteve and Adium and she serves on the advisory board of Johnson&Johnson; MT is a full-time employee and own stock in F. Hoffmann-La Roche Ltd; EB was a full-time employee in F. Hoffmann-La Roche Ltd; GK is a full-time employee and own stock in F. Hoffmann-La Roche Ltd; CQ-R is a full-time employee of Roche Diagnostics International Ltd, Rotkreuz, Switzerland; GK is a full-time employee of Roche Diagnostics GmbH, Penzberg, Germany; HZ has served at scientific advisory boards and/or as a consultant for Abbvie, Acumen, Alector, Alzinova, ALZpath, Amylyx, Annexon, Apellis, Artery Therapeutics, AZTherapies, Cognito Therapeutics, CogRx, Denali, Eisai, Enigma, LabCorp, Merck Sharp & Dohme, Merry Life, Nervgen, Novo Nordisk, Optoceutics, Passage Bio, Pinteon Therapeutics, Prothena, Quanterix, Red Abbey Labs, reMYND, Roche, Samumed, ScandiBio Therapeutics AB, Siemens Healthineers, Triplet Therapeutics, and Wave, has given lectures sponsored by Alzecure, BioArctic, Biogen, Cellectricon, Fujirebio, LabCorp, Lilly, Novo Nordisk, Oy Medix Biochemica AB, Roche, and WebMD, is a co-founder of Brain Biomarker Solutions in Gothenburg AB (BBS), which is a part of the GU Ventures Incubator Program, and is a shareholder of CERimmune Therapeutics (outside submitted work); KB has served as a consultant and at advisory boards for Abbvie, AC Immune, ALZPath, AriBio, Beckman-Coulter, BioArctic, Biogen, Eisai, Lilly, Moleac Pte. Ltd, Neurimmune, Novartis, Ono Pharma, Prothena, Quanterix, Roche Diagnostics, Sunbird Bio, Sanofi and Siemens Healthineers, has served at data monitoring committees for Julius Clinical and Novartis, has given lectures, produced educational materials and participated in educational programs for AC Immune, Biogen, Celdara Medical, Eisai and Roche Diagnostics, and is a co-founder of Brain Biomarker Solutions in Gothenburg AB (BBS), which is a part of the GU Ventures Incubator Program, outside the work presented in this paper; JDG receives research funding from Roche Diagnostics and GE Healthcare and has given lectures at symposia sponsored by Biogen and Philips; OG-R receives research funding from Roche Pharma; GS-B worked as a consultant for Roche Farma, S.A.

