## Supplementary Material for "Decreased Awareness of Cognitive Decline is Associated with Multimodal Alzheimer’s Disease Biomarkers in Cognitively Unimpaired Individuals"

§The complete list of collaborators of the ALFA study can be found in the acknowledgments section.

***Corresponding authors:**

David López-Martos, MSc
Barcelonaβeta Brain Research Center (BBRC), Pasqual Maragall Foundation.
Wellington 30, 08005, Barcelona, Spain


Gonzalo Sánchez-Benavides, PhD.
Barcelonaβeta Brain Research Center (BBRC), Pasqual Maragall Foundation.
Wellington 30, 08005, Barcelona, Spain


Oriol Grau-Rivera, MD, PhD
Barcelonaβeta Brain Research Center (BBRC), Pasqual Maragall Foundation.
Wellington 30, 08005, Barcelona, Spain


### Supplementary Material

**Supplementary Table 1.** Baseline Characteristics of the Participants with Episodic Memory Decline: Sensitivity Analysis

| | N (Sensitivity) | Episodic Memory Decline (n= 61) | Stable ACD (n= 36) | Decreased ACD (n= 25) | *p*-value |
|---|---|---|---|---|---|
| *Demographic* | | | | | |
| Age, mean (SD) | 61 (36 /25) | 62.13±5.23 | 61.24±5.06 | 63.41±5.30 | 0.112 |
| Sex [female], n (%) | 61 (36 /25) | 33 (54.1) | 16 (44.44) | 17 (68) | 0.120 |
| Education (years), mean (SD) | 61 (36 /25) | 13.34±3.66 | 13.83±3.73 | 12.64±3.51 | 0.213 |
| *Genetic* | | | | | |
| *APOE*-ε4 [carriership], n (%) | 61 (36 /25) | 35 (57.38) | 23 (63.89) | 12 (48) | 0.332 |
| *Biomarker* | | | | | |
| Plasma p-tau217 (pg/mL), mean (SD) | 61 (36 /25) | 0.1767±0.0937 | 0.1626±0.0896 | 0.1971±0.0975 | 0.160 |
| CSF p-tau181/Aβ42 (hybrid ratio), mean (SD) | 61 (36 /25) | 0.0200±0.0198 | 0.0181±0.0214 | 0.0229±0.0173 | 0.356 |
| CSF Aβ-positive (Aβ42/Aβ40), n (%) | 61 (36 /25) | 31 (50.82) | 16 (44.44) | 15 (60) | 0.350 |
| [$^{18}$F]flutemetamol PET (Centiloid units), mean (SD) | 51 (31 / 20) | 12.7650±23.2106 | 7.3007±19.1905 | 21.2346±26.6833 | **0.035** |
| *Clinical* | | | | | |
| mPACC, mean (SD) | 61 (36 /25) | -0.37±0.65 | -0.19±0.56 | -0.64±0.70 | **0.007** |
| MMSE, mean (SD) | 61 (36 /25) | 28.90±1.00 | 28.92±0.94 | 28.88±1.09 | 0.889 |
| Episodic Memory Composite, mean (SD) | 61 (36 /25) | -0.39±0.64 | -0.40±0.53 | -0.38±0.77 | 0.933 |
| SCD-Q (Memory), mean (SD) | 61 (36 /25) | 1.80±2.44 | 2.08±2.81 | 1.40±1.76 | 0.286 |
| Study Partner SCD-Q (Memory), mean (SD) | 61 (36 /25) | 0.89±1.32 | 0.72±1.09 | 1.12±1.59 | 0.250 |
| Study Partner A-IADL-Q, mean (SD) | 61 (36 /25) | 68.20±1.80 | 68.42±1.81 | 67.89±1.79 | 0.261 |

Results show mean and standard deviation (SD) for continuous variables, and number of observations (n) and percentage (%) for categorical variables [with reference level specified]. The first column corresponds to the sample of participants with episodic memory decline, the second and third columns correspond to the division by ACD groups. The previously validated CSF biomarker cut-off used to define CSF Aβ-positive in the ALFA+ cohort study was < 0.071 for the CSF Aβ42/40 ratio. Baseline characteristics between groups were evaluated using univariate ANOVAs for continuous variables and Chi-square tests for categorical variables. The mPACC was computed as a composite score derived from multiple cognitive domains, referenced to the Aβ-negative group, with higher scores indicating better cognitive performance. The MMSE captured global cognitive function, with higher scores indicating better performance. The Episodic Memory Composite was computed as z-score derived from the memory tests, referenced to the Aβ-negative group, with higher scores indicating better objective memory performance. The SCD-Q (memory domain) captured self-reported perception of memory difficulties, with higher scores reflecting greater subjective memory complaints; the Study Partner SCD-Q captured informant-reported perception of the participant's memory difficulties, with higher scores reflecting greater perceived complaints. The A-IADL-Q, completed by the study partner, assessed functional capacity in daily activities, with lower scores indicating greater functional difficulties.

**Supplementary Table 2.** Associations with Clinical Measurements: Sensitivity Analysis

| **Outcomes** | **n** | **Std. β (95% CI)** | ***p*-value** | **$R^2$** | **AIC** |
|---|---|---|---|---|---|
| mPACC | 61 (36 /25) | -0.591 (-1.093, -0.088) | **0.022** | 0.086 / 0.070 | 142.101 |
| Study Partner SCD-Q (Memory) | 61 (36 /25) | 0.309 (-0.210, 0.828) | 0.238 | 0.024 / 0.007 | 219.595 |
| Study Partner A-IADL-Q | 61 (36 /25) | -0.371 (-0.888, 0.145) | 0.155 | 0.034 / 0.018 | 247.409 |

Results from linear regression models indicating outcomes, number of observations (n; Stable SCD / Decreased ACD), standardized (Std.) β coefficients with 95% Confidence Intervals (CI), nominal p-values, R2 (i.e., nominal/adjusted proportion of explained variance), and Akaike Information Criteria (AIC) measurements. Clinical measures, mPACC, SCD-Q, and A-IADL-Q were determined using longitudinal complex SRB methods adjusted for age, sex, education, and time, entered as outcomes in independent models using the group classification (0: Stable ACD, 1: Decreased ACD) as the main predictor. Bold text indicates a significant association (p-value < 0.05).

**Supplementary Table 3.** Associations with Plasma and CSF Biomarkers: Sensitivity Analysis

| Outcomes | n | Std. β (95% CI) | *p*-value | $R^2$ | AIC |
|---|---|---|---|---|---|
| Plasma p-tau181 | 60 (36 / 24) | 0.491 (-0.025, 1.007) | 0.062 | 0.059 / 0.043 | -54.415 |
| Plasma p-tau181/Aβ42 | 60 (36 / 24) | 0.520 (0.006, 1.034) | **0.047** | 0.066 / 0.050 | -45.332 |
| Plasma p-tau217 | 61 (36 /25) | 0.403 (-0.111, 0.918) | 0.122 | 0.040 / 0.024 | -22.761 |
| CSF p-tau181 | 61 (36 /25) | 0.316 (-0.203, 0.835) | 0.228 | 0.025 / 0.008 | -18.960 |
| CSF p-tau181/Aβ42 | 61 (36 /25) | 0.431 (-0.082, 0.944) | 0.098 | 0.046 / 0.030 | 16.969 |
| CSF p-tau217 | 61 (36 /25) | 0.564 (0.059, 1.068) | **0.029** | 0.078 / 0.062 | 32.072 |

Results from linear regression models indicating outcomes, number of observations (n; Stable ACD / Decreased ACD), standardized (Std.) β coefficients with 95% Confidence Intervals (CI), nominal p-values, R2 (i.e., nominal/adjusted proportion of explained variance), and Akaike Information Criteria (AIC) measurements. Linear regression models used the group classification as main predictor of interest (0: Stable ACD, 1: Decreased ACD). Bold text indicates a significant association (p-value < 0.05).

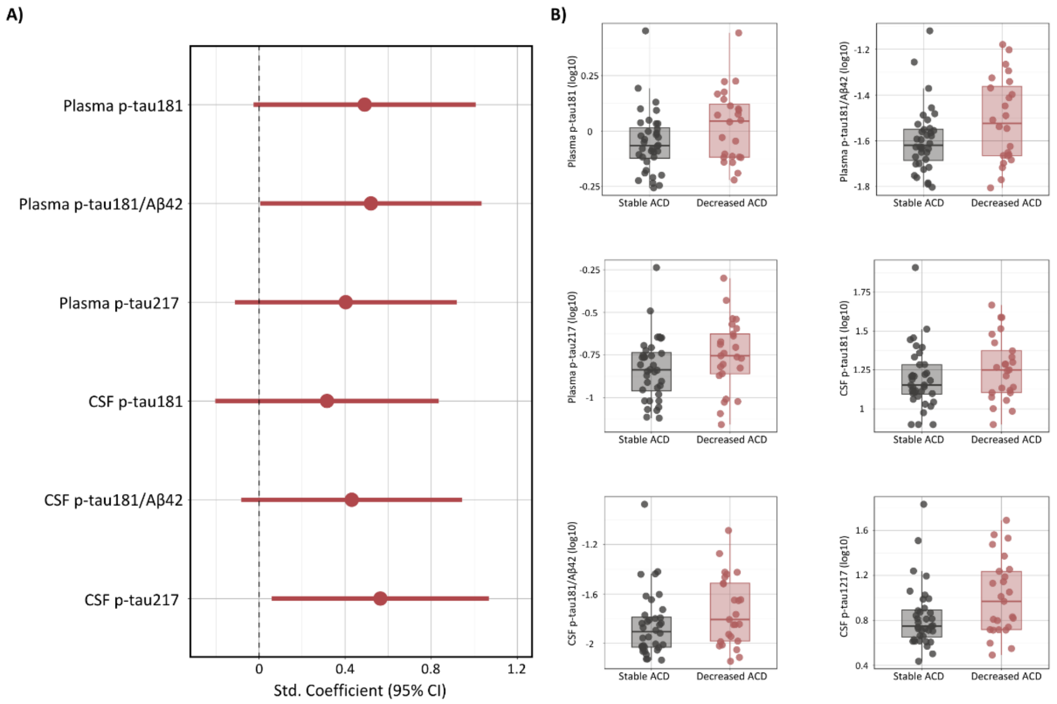


#### **Supplementary Figure 1.** Sensitivity Analysis: Associations of Decreased Awareness of Cognitive Decline with Plasma and CSF Biomarkers

A) Forest plot showing standardized regression coefficients and 95% confidence intervals for the effect of group (ACD classification; 0: Stable ACD, 1: Decreased ACD) on plasma and CSF biomarker outcomes from linear regression models adjusted for age and sex. B) Raw boxplots displaying the distribution of plasma and CSF biomarkers between groups.

**Supplementary Table 4.** Associations with PET Biomarkers: Sensitivity Analysis

| | | | Cluster-level | | Peak-level | | Peak MNI | | |
|---|---|---|---|---|---|---|---|---|---|
| **Outcomes** | **n** | **Anatomical locations** | **k** | ***p*-value** | **T** | ***p*-value** | **x** | **y** | **z** |
| Aβ PET | 34 (21 / 13) | | | | | | | | |
| | | Superior Frontal Right | 190 | 0.126 | 5.202 | **< 0.001** | 30 | 24 | 56 |
| | | Superior Temporal Right | 2864 | < 0.001 | 4.878 | **< 0.001** | 68 | -24 | 6 |
| | | Inferior Frontal Right | 2618 | < 0.001 | 4.377 | **< 0.001** | 56 | 34 | 4 |
| | | Middle Frontal Left | 1621 | < 0.001 | 4.352 | **< 0.001** | -42 | 36 | 28 |
| | | Supramarginal Left | 451 | 0.025 | 4.311 | **< 0.001** | -60 | -32 | 32 |
| | | Occipital Mid Left | 333 | 0.049 | 4.296 | **< 0.001** | -50 | -74 | 16 |
| | | Precentral Left | 1025 | 0.002 | 4.244 | **< 0.001** | -56 | -4 | 34 |
| | | Supplementary Motor Area Right | 316 | 0.055 | 4.030 | **< 0.001** | 8 | 20 | 62 |
| | | Superior Parietal Left | 246 | 0.085 | 3.778 | **< 0.001** | -20 | -62 | 60 |
| | | Middle Cingulate Right | 295 | 0.062 | 3.635 | **< 0.001** | 4 | 22 | 38 |
| | | Putamen Left | 595 | 0.012 | 3.592 | **0.001** | -22 | 4 | 2 |
| | | Paracentral Lobule Left | 100 | 0.259 | 3.473 | **0.001** | -4 | -28 | 58 |
| | | Posterior Cingulate Right | 1031 | 0.002 | 3.467 | **0.001** | 6 | -46 | 22 |
| | | Superior Frontal Left | 270 | 0.073 | 3.359 | **0.001** | -8 | 52 | 30 |
| | | Orbitofrontal Right | 236 | 0.091 | 3.160 | **0.002** | -2 | 62 | -18 |
| TAU PET | 23 (15 / 8) | | | | | | | | |
| | | Medial Temporal Lobe Left | 1931 | 0.001 | 4.656 | **< 0.001** | -20 | -26 | -22 |
| | | Middle Temporal Right | 3558 | < 0.001 | 4.587 | **< 0.001** | 50 | -70 | 12 |
| | | Insula Left | 167 | 0.263 | 3.902 | **< 0.001** | -38 | 0 | -6 |

Results from whole-brain voxel-wise linear regression models indicating outcomes, number of observations (n; Stable ACD / Decreased ACD), cluster-level anatomical location defined using 1mm3 resolution cluster labelling from the Automated Anatomical Labelling Atlas 3, cluster-level voxel size (k), cluster p-values, peak-level T-statistics, peak-level p-values, and peak-level coordinates according to the Montreal Neurological Institute (MNI) space. The voxel-wise neuroimaging models used the group classification as main predictor of interest (0: Stable ACD, 1: Decreased ACD). Bold text indicates a significant association (nominal p-values < 0.005 at the peak-level with a cluster-level threshold correction > 100 voxels).

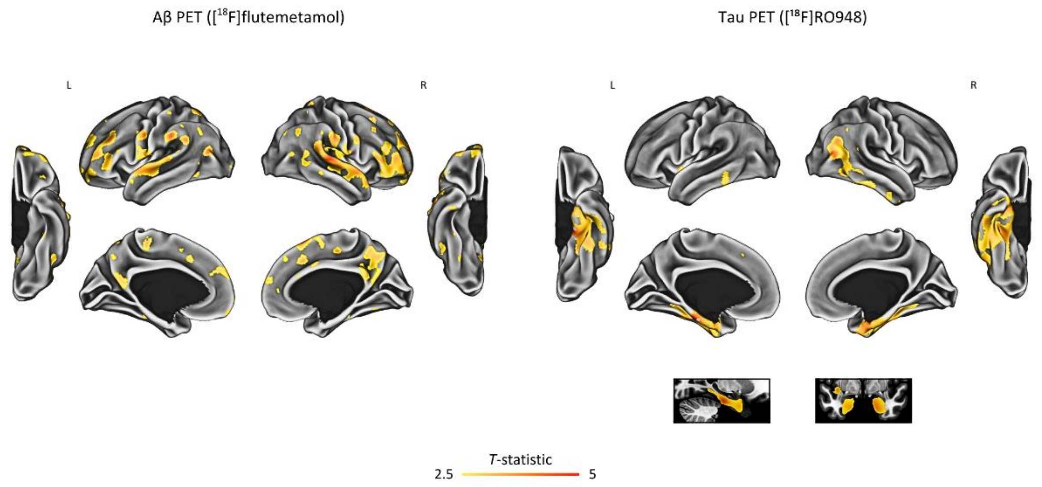


**Supplementary Figure 2.** Sensitivity Analysis: Associations of Decreased Awareness of Cognitive Decline with PET Biomarkers

Results from voxel-wise linear regression models showing the effect of group (0: Stable ACD, 1: Decreased ACD) on PET biomarker outcomes. The figure displays cortical surface-based projections following anatomical convention, where the left side of the image corresponds to the left hemisphere of the brain. Additionally, voxel-wise results are shown on T1-weighted anatomical sections. Color bars indicate *T*-statistics from vertex-wise projection into the cortical surface, with results shown at nominal *p*-values < 0.005 accounting for a cluster-size threshold of k > 100 voxels.
